# Assessment of a serological diagnostic kit of SARS-CoV-2 availble in Iran

**DOI:** 10.1101/2020.05.04.20090209

**Authors:** Hamid Reza Shamsollahi, Mostafa Amini, Shaban Alizadeh, Saharnaz Nedjat, Ali Akbari-Sari, Mehdi Rezaei, Seyed Farshad Allameh, Akbar Fotouhi, Masud Yunesian

## Abstract

**Background:** The SARS-CoV-2 epidemic broke out in December 2019 and now is characterized as a pandemic. The effective control of this infectious disease requires access to diagnostic techniques, both for case finding and epidemic size estimation. The molecular technique is routinely being used worldwide. Although it is the “standard” case detection and management method, it has its own shortcomings. Thus, some easy-to-use rapid serological tests were developed.

**Methods:** One hundred and fourteen positive RT-PCR-diagnosed patients were tested by VivaDiag Kit, a brand of rapid serological kits available in hospitals affiliated to Tehran University of Medical Sciences (TUMS), Tehran, Iran. Frozen serum specimens taken from healthy people in summer and autumn 2019, were also tested as negative controls.

**Results:** The test sensitivity was 47.9% (95% confidence interval [CI]: 38.8-56.9) for IgM and 47.0% (95% CI: 38.0-56.0) for IgG. There was no difference between IgG and IgM seropositivity except in one case. Specificity was calculated as 99.0% (95% CI: 96.4-99.9) for IgM and of 100.0% (95% CI: 0.98.2-100.0) for IgG. Sensitivity was higher in men and older participants.

**Conclusion:** This test can be used for epidemiological investigations especially for the estimation of the level of infection in the community, after it is properly corrected for sensitivity and specificity. The low sensitivity could be attributed to the technical limitation of the kits or low levels of antibodies after infection. The different sensitivity in age and sex groups supports the hypothesis that different people show different immune responses to this virus.

## Introduction

The world health organization (WHO) has recommended some strategies to control the transmission of SARS-CoV-2 in the general population. One of the most important of these strategies is active case finding and isolation, followed by contact tracing and social distancing (physical distancing) (1). The key factor contributing to the successful isolation of infected cases is the early diagnosis of infection with high sensitivity, especially in asymptomatic and/or preclinical cases. As reported in a study at Columbia University Irving Medical Center (CUIMC) (New York, USA), 29 out of 33 tested positive pregnant women (based on nasopharyngeal PCR test) admitted to a hospital were asymptomatic cases (2). Hence, gaining access to appropriate diagnostic tests is crucial for a successful strategy. The reverse transcription-polymerase chain reaction (RT-PCR) is the proposed method for diagnostic purposes for those who are suspected of COVID-19 infection (3). Nevertheless, there are some problems with the widespread usage of this test. For a successful RT-PCR test, an appropriate nasopharyngeal sampling is required at the onset of virus shedding. This is difficult and may endanger healthcare personnel in some cases, requiring well-trained health workers as well as suitable transport media for taking the sample and sending it to the laboratory. The RT-PCR itself needs trained virologists and appropriate devices for genomic DNA (gDNA) extraction and performing the test. Besides, it takes 1 to 2 days for the result to be ready. Moreover, the RT-PCR cannot detect infections before or after the onset of virus shedding (i.e. in pre-clinical cases and recovered patients). In addition, since the RT-PCR is an expensive test, many companies are trying to develop serological test kits to facilitate the diagnosis of COVID-19 based on the detection of IgM and IgG. Furthermore, *rapid test* kits are commonly developed and used by some institutes for this purpose, which can return results within 15 minutes, according to the developers (4). Thanks to their advantages (e.g., cost-effectiveness, simplicity, and rapidness of the results), using rapid serological tests seems tempting for case detection over molecular tests. They can also be used in epidemiological studies for the detection of infection history in recovered patients. There are, however, some technical problems with using these tests too. As WHO has mentioned in its technical document, these tests are only applicable for epidemiological purposes and should not be used in clinical settings right now(3), mainly due to the incubation period between infectivity and seropositivity. Even for epidemiological purposes, it is crucial to know the exact sensitivity and specificity of these tests being evaluated by independent researchers, before application in large-scale studies on the general population. The sensitivity and specificity of one of these tests that are commonly used in some research settings in Iran were evaluated in this study. This test was developed to detect IgM and IgG antibodies in whole blood, serum, and plasma of patients. The developer claimed a very high sensitivity (94.4%) and specificity (100%) for the test within 11-24 days after infection.

## Material and Methods

The test kit used in this research for the detection of COVID-19 antibodies is manufactured by VivaChek Inc., based in China, under the commercial name “VivaDiag” lot No. E2002002. A sample kit was donated to Tehran University of Medical Sciences (TUMS) by the Ministry of Health and Medical Education (MOHME). It reports the presence or absence of both IgM and IgG antibodies in samples qualitatively. IgM usually indicates active or recently recovered infection and IgG can detect infection history several months or even years after recovery (5). RT-PCR-confirmed COVID-19 cases in several hospitals affiliated to TUMS were enrolled as *true positive* cases. Likewise, a random sample of frozen sera of healthy people participating in TUMS Employees COHORT (TEC) study that was taken in summer and autumn 2019 (months before reporting the first case of COVID-19 by China) was used as the *true negative* sample. Based on kit instructions, 10 μL blood was put in the kit specimen well. Then, two to three drops of buffer solution were added based on kit instructions. Then, the kit was left for 15 minutes, after which the appeared result for IgM and IgG were recorded. The same process was repeated for true negative frozen serum specimens after thawing. Age, sex, time of first sign/symptom, time of serological test, and some other demographic characteristics were also recorded through interviews using a researcher-made checklist. For true negative samples, demographic variables were acquired from TEC records. The sensitivity of the kit was calculated by dividing the number of positive test results for IgG, IgM, or each of these markers by total RT-PCR confirmed COVID-19 cases. Its specificity was calculated separately for IgG and IgM antibodies by dividing the number of negative test results by the total true negative samples. A 95% confidence interval for both sensitivity and specificity was calculated based on the binomial distribution. The study has been approved by TUMS Ethics Committee (approval No. IR.TUMS.VCR.REC. 1399.045).

## Results

A total of 114 PCR-confirmed patients and 198 negative sera were tested. The average age was 44.0 (±12.1) for the first group and 39.2 (±8.0) for the second group. Moreover, it took 5-53 days from the appearance of the first sign or symptom to testing.

The sensitivity and specificity of the applied kit for IgM and IgG are shown in Table 1.

**Table 1.** - The sensitivity and specificity of VivaDiag Test Kit

| Antibody | Total Tests (n) | True Responses (n) | Point Estimate (%) | 95% CI |
| --- | --- | --- | --- | --- |
| **Sensitivity** |  |  |  |  |
| IgM | 114 | 54 | 47.4 | 37.9 - 56.9 |
| IgG | 114 | 53 | 46.5 | 37.1 - 56.1 |
| IgM or IgG | 114 | 54 | 47.4 | 37.9 - 56.9 |
| **Specificity** |  |  |  |  |
| IgM | 198 | 196 | 99.0 | 96.4 - 99.9 |
| IgG | 198 | 198 | 100 | 98.2 - 100 |

CI: confidence interval

The results were also classified into three groups based on the time it takes from the appearance of the first signs or symptoms of the illness and serological test, as shown in Table 2.

**Table 2.** - The sensitivity of VivaDiag Test Kit at different intervals from the onset of symptoms to testing

| Interval (Day) | IgM (%)<br>(95% CI) | IgG (%)<br>(95% CI) | IgG or IgM (%)<br>(95% CI) |
| --- | --- | --- | --- |
| 0-19 | 41.9<br>(24.6-60.9) | 38.7<br>(21.9-57.8) | 41.9<br>(24.6-60.9) |
| 20-39 | 52.3<br>(39.5-64.9) | 52.3<br>(39.5-64.9) | 52.3<br>(39.5-64.9) |
| $\geq 40$ | 38.9<br>(17.3-64.3) | 38.9<br>(17.3-64.3) | 38.9<br>(17.3-64.3) |
| p-value | 0.467 | 0.358 | 0.467 |
| p-value for trend | 0.964 | 0.762 | 0.964 |

CI: confidence interval

Different age groups and the two sexes were also compared in terms of the detection rate. Men and those older than 50 had higher detection rates (Table 3)

**Table 3.** - The sensitivity of the VivaDiag Test Kit concerning sex and different age groups

| **Variable** | **IgM (%)**<br>**(95% CI)** | **IgG (%)**<br>**(95% CI)** | **IgG or IgM (%)**<br>**(95% CI)** |
| --- | --- | --- | --- |
| **Sex** |  |  |  |
| Male | 63.0<br>(48.8-75.7) | 63.0<br>(48.8-75.7) | 63.0<br>(48.8-75.7) |
| Female | 33.3<br>(21.7-46.7) | 31.7<br>(20.2-45.0) | 33.3<br>(21.7-46.7) |
| p-value | 0.001 | 0.002 | 0.001 |
| **Age Group** |  |  |  |
| <50 years | 40.7<br>(30.0-52.2) | 39.5<br>(28.8-51.0) | 40.7<br>(30.0-52.2) |
| ≥50 years | 63.6<br>(45.1-79.6) | 63.6<br>(45.1-79.6) | 63.6<br>(45.1-79.6) |
| p-value | 0.026 | 0.019 | 0.026 |

## Discussion

This study evaluated VivaDiag, a COVID-19 commercial rapid test kit, to determine its capability in diagnosing PCR-confirmed COVID-19 patients rather than truly infected people. As claimed by the manufacturer, the kit was expected to has a sensitivity of 94.4% and 100% for IgM and IgG, 11-24 days after infection, respectively. According to the results of this study, the sensitivity ranges from 38.9 to 52.3 in different time intervals after infection, with no statistically significant difference between the kinds of antibody (IgG and IgM) and between different intervals between disease onset and antibody testing.

Other researchers have reported lower sensitivity levels for this kit, or other commercial kits. *Cassaniti et al*. (2020), for example, reported an 18.4% sensitivity for this kit based on IgM or IgG for a sample of 50 confirmed cases in Italy. They were not adjusted, however, for the time interval between disease onset and testing time (6). In a study conducted in Santa Clara, California, *Bendavid et al*. (2020) also reported a 67.65% sensitivity for IgM in Premier Biotech (USA) kits as opposed to the 91.8% sensitivity claimed by the manufacturer (7).

It is difficult to justify the apparent difference between the sensitivity of the same kit in different studies or subgroups of people, as the sensitivity and specificity of the tests are expected to be independent of the population being tested, from a technical point of view. This may be due to the fact that different people respond differently to virus exposure, as a hypothesis. This hypothesis was strengthened by different sensitivities observed in different age and sex groups, which may be due to different immune responses to infection in these groups. These different immune responses may also be attributed to higher case fatality rates in men and older people. The specificity of both antibodies was claimed to be 100%. The results of this study also confirmed a high specificity (99% for IgM and 100% for IgG) with 96.4 and 98.2 as the lower bound of the 95% confidence interval, respectively. Based on our findings, the positive cases detected by this kit are reliable. Moreover, using this test for epidemiological purposes will be useful, provided that the actual prevalence of infection is not less than 2% (the upper bound of the confidence interval for false-positive results).

Although rapid serological tests cannot be regarded as useful in “effective screening,” they can be applied in epidemiological studies if the exact (narrow) estimate of test sensitivity and specificity is available. By knowing the exact number of infected cases, we can calculate the infection fatality rate of infectious diseases. Although *Bendavid et al*. (2020) announced a 50-85-fold under-ascertainment rate in their paper, this finding is highly associated with the test performance, especially to the test specificity (7).

## Conclusion

The evaluated rapid kit lacks acceptable sensitivity to be able to detect infected people properly. Positive results, however, can yield a good estimate of the prevalence of infection and may help us get a better idea of infection fatality rate, after being corrected for incomplete sensitivity and specificity. COVID-19 is a novel viral infection and the serological tests for its detection are still in their infancy. Any recommendation on general uses of these kits for diagnostic or even epidemiological purposes entails the evaluation of their accuracy using a reasonable sample size of confirmed positive and negative samples by independent researchers in different populations. Different sensitivities in different age and sex groups could be ascribed to different immune system responses and antibody titers among these groups.

## Data Availability

All the data is available in the passage.

## Ethical Statement

This research has been approved by TUMS Research Ethics Committee. All participants completed the questionnaires after being informed of research objectives and getting verbal consent from them.

## Acknowledgment

The authors are grateful to the deputy secretary of treatment affairs at Tehran University of Medical Sciences (TUMS), Tehran, Iran for his contribution to testing and filling out the questionnaire. We also thank MS. Masoumeh Barati, MS. Elaheh Kimiaei, MS. Asma Javadi Rad, MS. Zahra Kashani, Mr. Alireza Ranjbarian, and MS. Anahita Sedaghat for their kind contribution in data collection.

